# Developing a Phenotype Risk Score for Tic Disorders in a Large, Clinical Biobank

**DOI:** 10.1101/2023.02.21.23286253

**Authors:** Tyne W. Miller-Fleming, Annmarie Allos, Emily Gantz, Dongmei Yu, David A. Isaacs, Carol A. Mathews, Jeremiah M. Scharf, Lea K. Davis

## Abstract

**Importance:** Tics are a common feature of early-onset neurodevelopmental disorders, characterized by involuntary and repetitive movements or sounds. Despite affecting up to 2% of young children and having a genetic contribution, the underlying causes remain poorly understood, likely due to the complex phenotypic and genetic heterogeneity among affected individuals.

**Objective:** In this study, we leverage dense phenotype information from electronic health records to identify the disease features associated with tic disorders within the context of a clinical biobank. These disease features are then used to generate a phenotype risk score for tic disorder.

**Design:** Using de-identified electronic health records from a tertiary care center, we extracted individuals with tic disorder diagnosis codes. We performed a phenome-wide association study to identify the features enriched in tic cases versus controls (N=1,406 and 7,030; respectively). These disease features were then used to generate a phenotype risk score for tic disorder, which was applied across an independent set of 90,051 individuals. A previously curated set of tic disorder cases from an electronic health record algorithm followed by clinician chart review was used to validate the tic disorder phenotype risk score.

**Main Outcomes and Measures:** Phenotypic patterns associated with a tic disorder diagnosis in the electronic health record.

**Results:** Our tic disorder phenome-wide association study revealed 69 significantly associated phenotypes, predominantly neuropsychiatric conditions, including obsessive compulsive disorder, attention-deficit hyperactivity disorder, autism, and anxiety. The phenotype risk score constructed from these 69 phenotypes in an independent population was significantly higher among clinician-validated tic cases versus non-cases.

**Conclusions and Relevance:** Our findings provide support for the use of large-scale medical databases to better understand phenotypically complex diseases, such as tic disorders. The tic disorder phenotype risk score provides a quantitative measure of disease risk that can be leveraged for the assignment of individuals in case-control studies or for additional downstream analyses.

**Key Points:** *Question:* Can clinical features within the electronic medical records of patients with tic disorders be used to generate a quantitative risk score that can identify other individuals at high probability of tic disorders?

*Findings:* In this phenome-wide association study using data from electronic health records, we identify the medical phenotypes associated with a tic disorder diagnosis. We then use the resulting 69 significantly associated phenotypes, which include several neuropsychiatric comorbidities, to generate a tic disorder phenotype risk score in an independent population and validate this score with clinician-validated tic cases.

*Meaning:* The tic disorder phenotype risk score provides a computational method of evaluating and distilling the comorbidity patterns that characterize tic disorders (independent of tic diagnosis status) and may help improve downstream analyses by distinguishing between individuals that should be categorized as cases or controls for tic disorder population studies.

## Introduction

Tic disorders (TD) are the most common movement disorder in children and are characterized by sudden and recurrent movements and/or vocalizations^1–4^. While many tic symptoms resolve within a year, persistent TD can cause disruptions to daily life and may have long-term effects on an individual’s social, physical, and mental health^5–7^. TD is highly comorbid with several other psychiatric conditions, including obsessive-compulsive disorder, attention-deficit hyperactivity disorder, and autism, among others^8–17^. One study found that 86% of individuals diagnosed with Tourette syndrome will be diagnosed with one additional psychiatric disorder and up to 58% of Tourette syndrome patients will be diagnosed with two or more additional psychiatric disorders in their lifetime^13^. This phenotypic heterogeneity across affected individuals complicates the diagnosis and treatment of patients with Tourette syndrome and other TDs.

Electronic health records (EHRs) have become a useful resource for studying disease outcomes and comorbidities^18^. EHR systems often date back several years to decades and document a wide range of phenotype information, including diagnosis and billing codes, clinician notes, medical histories, lab results, medications, and procedural codes. Additionally, EHR systems allow rapid scaling for genome-wide association studies. The inclusion of large samples across diverse backgrounds and disease groups without requiring the time and funds needed to recruit large cohorts allows a massive advantage for genomic studies of rare or underdiagnosed phenotypes like TD. Furthermore, in the case of phenotypically complex diseases, such as TD, EHRs can provide dense phenotype information spanning before and after disease diagnosis.

The recent use of phenotype risk scores (PheRS) calculated from medical records has successfully identified patients that exhibit overlapping features of disease. Similar to genetic risk scores, a discovery cohort is used to extract the phenotypic features that define a disease in the medical records. These features are then evaluated within an independent target population and each individual is scored based on the number of disease features they exhibit. Individuals with a high PheRS are phenotypically similar to the diagnosed individuals of the discovery dataset, whereas individuals with a low PheRS share little or no phenome of the disease. The PheRS method was initially developed to identify patients with shared features of Mendelian diseases within the medical record database but has recently been applied to neuropsychiatric conditions, including major depressive disorder, generalized anxiety disorder, and posttraumatic stress disorder^19–22^. This method condenses the unique medical phenome, which in this case aligns with a TD diagnosis, into a single quantitative measure that can then be used for downstream analyses. Additionally, because this method relies solely on the collection of diagnosis codes within the medical record, it is a powerful tool for indexing liability for TD, given that tics are a common comorbidity but are uncommonly explicitly coded or charted in a medical record.

In this study we leverage de-identified medical records from the Vanderbilt EHR system to identify the phenotypic correlates of TD. These features were then used to generate and deploy a tic disorder PheRS within an independent cohort. The TD PheRS is significantly higher for clinically-validated tic patients versus non-diagnosed individuals, supporting the utility of medical records for evaluating phenotypically complex diseases and providing a framework for using the PheRS as a tool for phenome-wide investigations.

## Methods

### Sample Description

For the tic disorder phenome-wide association study (PheWAS), TD cases and controls within the EHR (Vanderbilt synthetic derivative, N=3.6 million) were identified by ICD-9 and ICD-10 (International Classification of diseases, Ninth/Tenth Revision) billing codes using the criteria outlined in Supplemental Methods (**Supplemental Table 1**). 1,406 individuals with TD ICD codes and 7,030 age and sex-matched controls (absent of TD ICD codes) were included in the analysis. The tic disorder PheRS was deployed in the independent BioVU population (see Supplemental Methods, N=90,031).

### Phenome-wide association study for TD status

PheWAS was performed to identify the medical phenotypes enriched in individuals with TD ICD codes (hereafter referred to as “diagnosed TD patients”). Diagnosed TD patients (N=1,406) and matched unscreened patients (N=7,030) were assigned a one or a zero status (exposure variable), respectively, and logistic regressions were performed across 676 phenotypes using the PheWAS package in R (version 0.99.5-2 and 3.6.0, respectively)^23,24^. The case/control outcome variables were defined by phecodes mapped from ICD9/10 billing codes across all medical diagnoses as previously described^25^. We required each individual to have at least 2 instances of an ICD9/10 within the medical record to be considered a case for any outcome. A minimum of 20 cases was required to test each phenotype. Covariates included sex, current age, median age within the medical record, EHR-reported race, EHR-reported ethnicity, and number of visits to the medical center. The 69 phenotypes with a p-value below the Bonferroni-corrected threshold *(P=*7.396 × 10^−5^, 0.05/676) were considered significant.

### PheWAS phenotype group enrichment

The phecodes analyzed in the TD PheWAS were mapped from ICD9/10 billing codes and were grouped into 17 independent phenotype groups. The hypergeometric test was performed to identify phenotype groups that were over- or under-represented in the 69 significant PheWAS results. Phenotype groups with a p-value below the Bonferroni-corrected threshold *(P=*0.00294, 0.05/17) were considered significantly over- or under-represented. The fold-enrichment or fold-depletion was reported for each phenotype group in **Figure 3B**.

### TD phenotype risk score construction

As described above, we performed a PheWAS on the presence/absence of TD diagnosis and identified 69 phenotypes significantly enriched or depleted in the TD diagnosed individuals. These 69 phenotypes were then used to construct the TD PheRS in the independent, genotyped BioVU population, or target sample. The following equation was used as previously described:

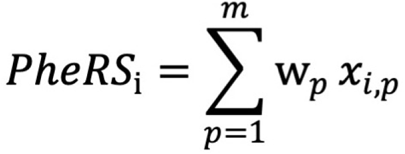

where *x*_*i,p*_ is equal to 1 if individual *i* has phenotype *p* or 0 if the phenotype is not present in the individual’s health records, and *w*_*p*_ refers to the weight of phenotype *p*. The weights for each phenotype are equal to the effect size estimate of each phenotype from the initial PheWAS of the discovery sample. Therefore, phenotypes with the strongest association to TD status will contribute the most to the PheRS, whereas phenotypes with weaker associations will contribute less. **Supplemental Table 8** lists the 69 contributing phenotypes with their respective weights. The PheRS was then calculated in 90,051 BioVU individuals (**Supplemental Table 7**).

## Results

### PheWAS Identifies the Complex Clinical Phenome of Tic Disorder Patients

To uncover the phenotypic correlates of tic disorders (TD) within a database of de-identified electronic health records (EHR), we performed a PheWAS of TD diagnosis presence/absence (**Figure 1, Supplemental Table 1**). As expected for an early-onset disorder with a male-bias, our EHR-derived TD cases (and matched controls) are relatively young (average age 24.22/24.25 years old, average median age of medical record 15.40/13.95 years old, and average age at first ICD code 11.39/9.02 years old in cases/controls) and predominantly male (72.48%/72.59% male in cases/controls). Consistent with the demographic characteristics of the EHR at Vanderbilt, TD cases and controls are significantly skewed for individuals with EHR-reported race and ethnicity as white and non-Hispanic (77.67%/75.02% white in TD present/absent and 84.57%/89.20% non-Hispanic in TD present/absent) (**Table 1**). We identified 69 phenotypes significantly associated with TD diagnosis after Bonferroni-correction (*P* < 7.396 × 10^−5^) (**Figure 2, Supplemental Table 2**). Included among the top associations are *tic disorders* and *tics of organic origin*, both of which are significantly associated with the TD case label (*P* = 4.18 × 10^−65^; β, 9.94; SE, 0.58 and *P* = 1.40 × 10^−42^; β, 6.92; SE, 0.51, respectively).

**Figure 1:**
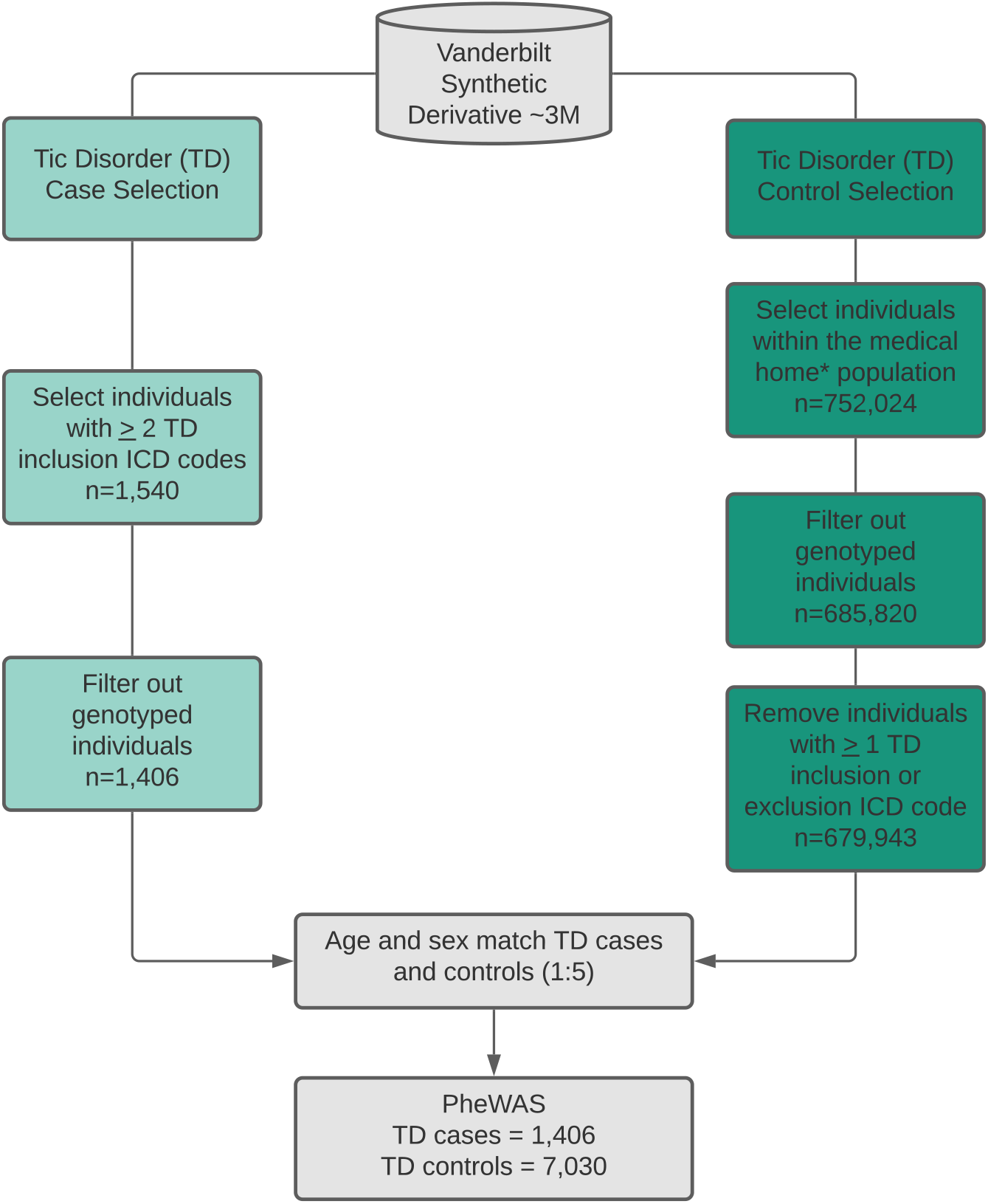
Extraction of tic disorder (TD) cases and controls from electronic health records (EHR). Tic disorder cases were identified by selecting individuals with 2 instances of the TD inclusion phenotypes. TD cases were restricted to the non-genotyped population in the EHR. TD controls were matched to TD cases after filtering out the genotyped individuals, those that did not belong to the medical home, and those with at least 1 inclusion or exclusion ICD code for TD. Medical home individuals are those that have visited a Vanderbilt clinic at least 5 times over a three-year period. Matching of cases and controls was performed at a 1:5 ratio, respectively, based on age and sex. ICD9/10 billing codes used as inclusion/exclusion criteria for TD cases and controls are listed in Supplemental Table 1.

**Table 1.**
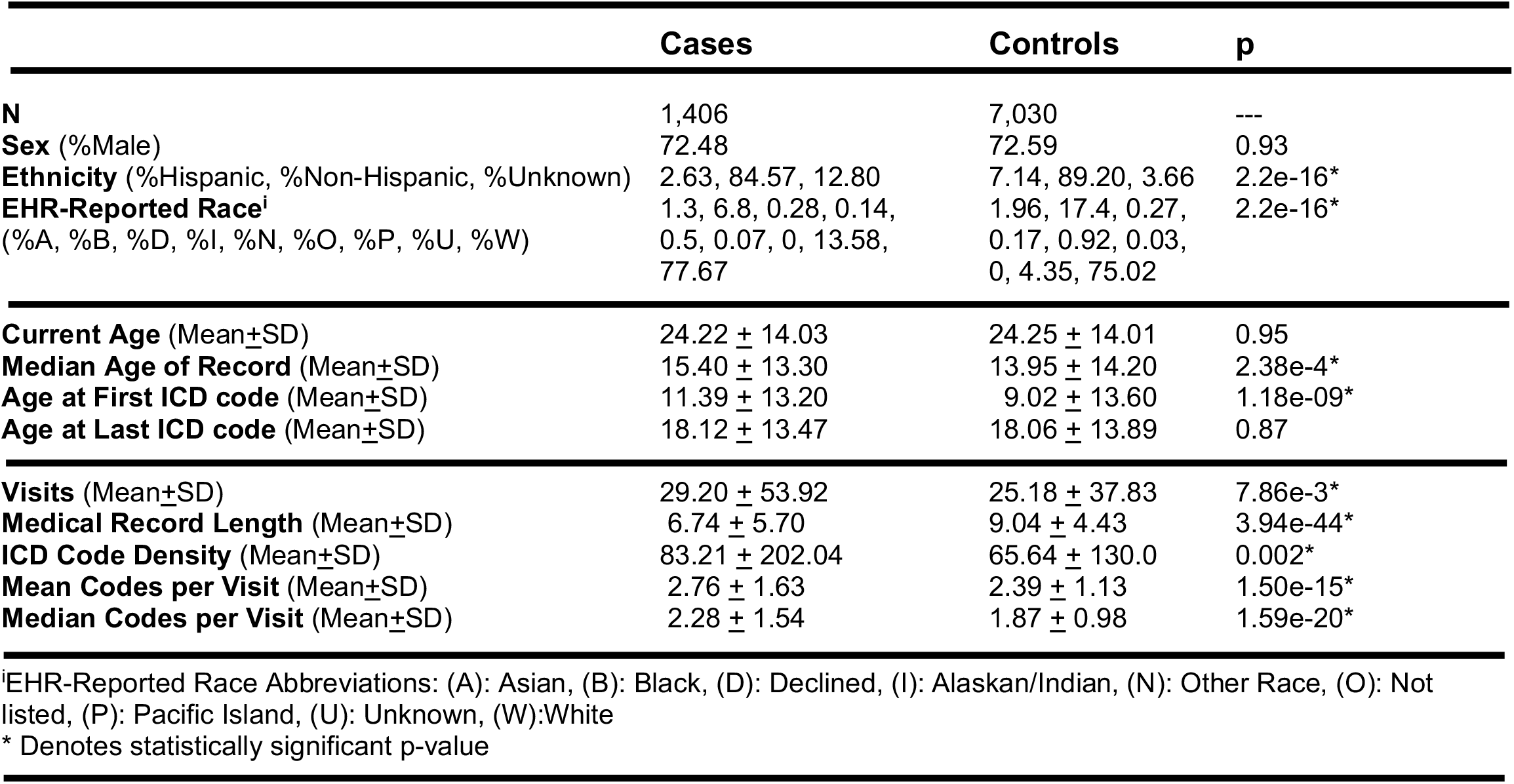
Demographics for Tic Disorder PheWAS. **Tic Disorder (TD) case-control demographics.** Tic disorder (TD) cases (N = 1,406) were identified using billing codes within the electronic health records. Tic disorder controls (N = 7,030) were matched to cases based on current age and sex. Independent-samples t-test or chi-squared analyses were performed on all measures to determine if there was a statistically significant difference between cases and controls.

**Figure 2:**
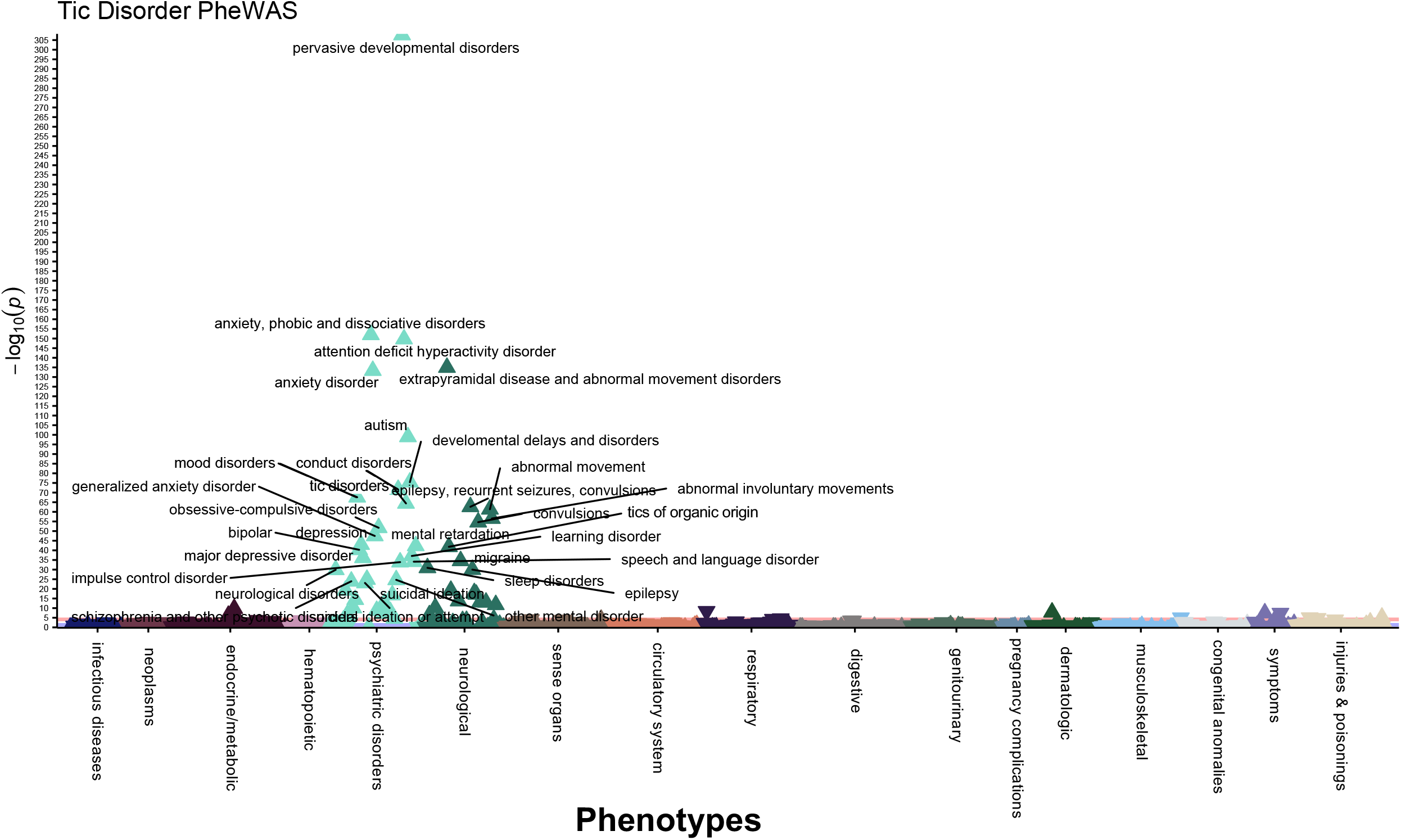
Tic disorder diagnosis PheWAS identifies phenotypic effects across the medical phenome. 676 phenotypes within the electronic health records were tested for enrichment in individuals with ICD9/10 diagnosis codes for TD (N=1,406) compared to age and sex-matched controls (N=7,030). Phenotypes were defined as phecodes mapped from ICD9/10 billing codes. Logistic regressions were performed for each phenotype, individuals were required to have 2 instances of a phenotype to be considered a case and at least 20 cases were required for testing each phenotype. 69 phenotypes were significantly enriched in the TD cases after Bonferroni-correction *(P =* 7.396 × 10^−5^, 0.05/676 number of tests). Phenotypes *with P* ≤ 5.0 × 10^−20^ are annotated on the Manhattan plot. Covariates included: current age, sex, EHR-reported race, EHR-reported ethnicity, median age of medical record, and number of visits to medical center.

**Figure 3:**
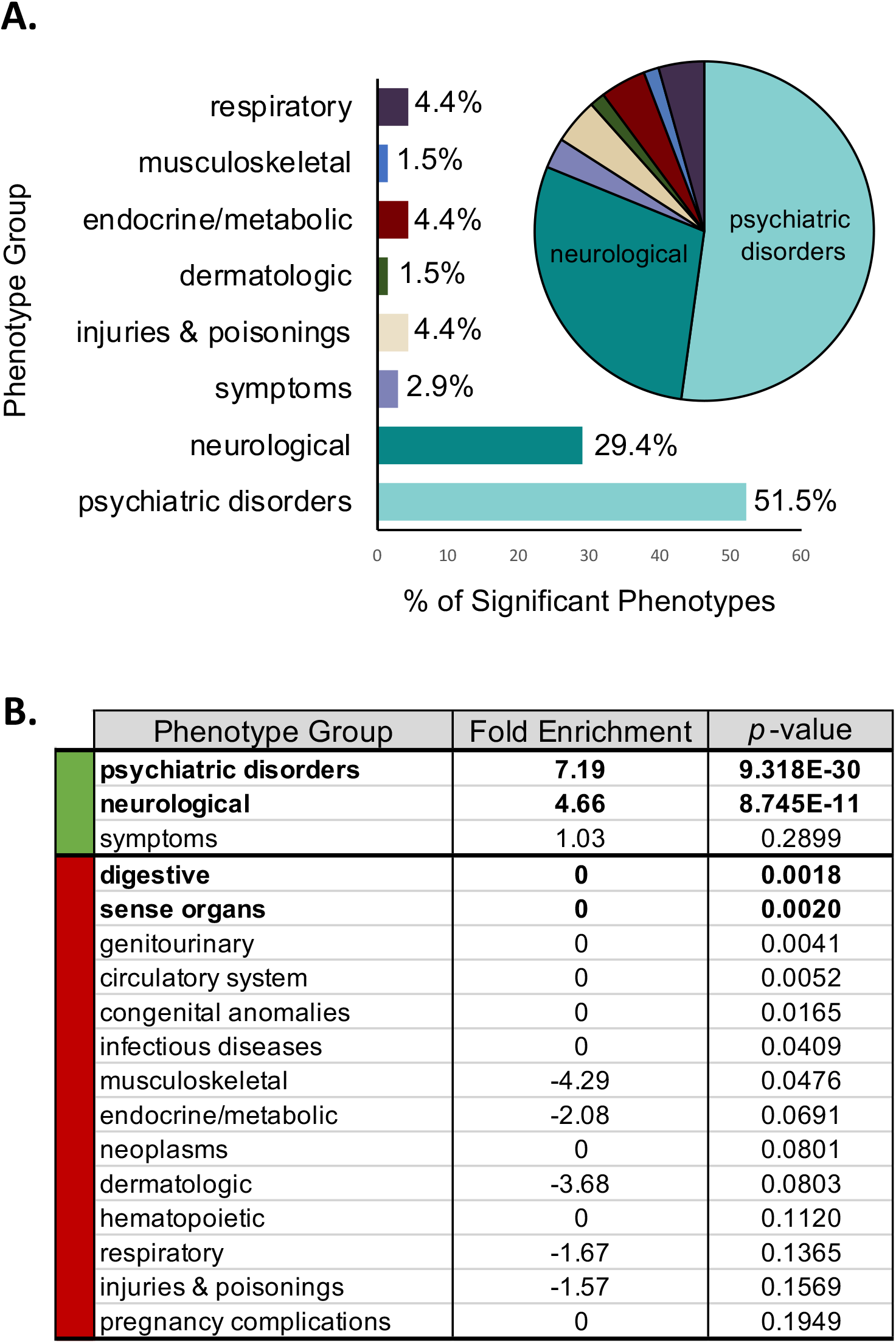
Tic disorder PheWAS is enriched for psychiatric disorders and neurological phenotypes. A. 69 phenotypes were significantly enriched or depleted within the tic disorder cases versus controls. The bar graph and pie chart show the proportions of these 69 phenotypes across 17 phenotype groups. Over 80% of the phenotypes enriched in the tic disorder individuals belong to the psychiatric disorders or neurological phenotype groups. B. The phenotype groups for psychiatric disorders and neurological phenotypes are significantly enriched in the tic disorder individuals (7.19 and 4.66 fold enrichment, respectively, *P* < 0.00294 calculated by the hypergeometric test). Digestive and phenotypes within the sense organs were significantly depleted within the tic disorder cases compared to controls (*P* < 0.00294 calculated by the hypergeometric test).

### Psychiatric and neurological conditions are highly comorbid in TD patients

As expected, in addition to tic-specific phenotypes, the top associations from the TD PheWAS included known comorbidities, such as: *anxiety disorders* (*P* = 1.12 × 10^−152^; β, 2.38; SE, 0.09)^13^, *attention deficit hyperactivity disorder (P =* 1.84 × 10^−150^; β, 3.43; SE, 0.13)^12^, *autism* (*P* = 1.17 × 10^−99^; β, 3.57; SE, 0.17)^15^, *developmental delays and disorders (P =* 3.99 × 10^−76^; β, 2.98; SE, 0.16), *conduct disorders (P =* 2.57 × 10^−72^; β, 3.41; SE, 0.19)^8^, *mood disorders (P =* 2.57 × 10^−68^; β, 1.83; SE, 0.10), *obsessive compulsive disorders (P =* 1.86 × 10^−52^; β, 4.29; SE, 0.28)^16^, *depression (P =* 9.46 × 10^−44^; β, 1.73; SE, 0.13)^26^, *migraine (P =* 2.70 × 10^−35^; β, 1.73; SE, 0.14)^27^, and *sleep disorders (P =* 1.19 × 10^−31^; β, 1.39; SE, 0.12)^28,29^. The most significantly associated phenotype was *pervasive developmental disorders (P <* 5 × 10^−324^; β, 4.80; SE, 0.11), a broad diagnosis that encompasses multiple disorders of impaired communication and socialization skills, including: autistic disorders, Asperger’s syndrome, Rett’s disorder, and childhood disintegrative disorder. This result is consistent with previous studies that found 1 in 22 individuals with Tourette Syndrome had comorbid pervasive developmental disorder (PDD), although the term PDD has fallen out of clinical use since being replaced by the term autism spectrum disorder in the DSM-5^1,30^. In the EHR, the phecodes for attention deficit hyperactivity disorder (313.1), tic disorders (313.2), and autism (313.3) are rolled into the PDD phecode (313), which explains our finding that 83.36% of the EHR-defined TD cases also have a diagnosis for PDD (**Supplemental Table 3**).

The TD label was enriched for several other neuropsychiatric phenotypes, such as: *bipolar disorder (P =* 5.11 × 10^−41^; β, 2.57; SE, 0.19), *suicidal ideation* (*P* = 1.38 × 10^−25^; β, 2.02; SE, 0.19), *schizophrenia and other psychotic disorders (P =* 1.16 × 10^−24^; β, 2.61; SE, 0.25), *transient alteration of awareness (P =* 6.33 × 10^−20^; β, 2.16; SE, 0.24), *adjustment reaction* (*P* = 1.52 × 10^−17^; β, 1.46; SE, 0.17), *personality disorders (P =* 1.89 × 10^−13^; β, 2.90; SE, 0.39), *posttraumatic stress disorder (P =* 6.93 × 10^−8^; β, 1.32; SE, 0.24), and *eating disorder* (*P* = 2.37 × 10^−5^; β, 1.59; SE, 0.38) (**Supplemental Table 3**). Among EHR-derived TD labels, 85.6% have at least one additional neuropsychiatric disorder diagnosis and 51.9% have two or more additional neuropsychiatric diagnoses, compared to 21.2% and 16.8%, respectively, within TD controls. These differences are statistically significant *(P* < 1 × 10^−4^, Fisher’s Exact Test) and are comparable to previous findings of psychiatric comorbidities within individuals with tic disorders^13^. The proportion of control individuals with at least one neuropsychiatric diagnosis is higher than expected, likely because of ascertainment bias of young individuals within the EHR, which tend to be enriched for neuropsychiatric phenotypes. Because our EHR database contains longitudinal phenotype information on the TD cases, we examined whether tic diagnoses are more commonly made prior to or after other psychiatric diagnoses. Greater than 64% of the TD cases were diagnosed with a tic disorder before receiving any additional neuropsychiatric diagnoses (**Supplemental Table 4**). These findings are consistent with the early onset nature of tic disorders and may provide clinicians with critical information regarding the disorders that tic patients are at greatest risk of developing later in life.

The 676 phenotypes assessed in our TD PheWAS are binned across 17 distinct phenotype groups, such as psychiatric disorders, neurological phenotypes, neoplasms, etc. To determine whether a specific phenotype group was over-represented among the 69 significant results, we used a hypergeometric test. Over 80% of the phenotypes enriched in TD cases map to psychiatric and neurological disorders (7.19-fold enrichment, *P* = 9.318 × 10^−30^ and 4.66-fold enrichment, *P* = 8.745 × 10^−11^, respectively) (**Figure 3A** and **Figure 3B**). The remaining significantly enriched phenotypes groups included: respiratory (4.4%), endocrine/metabolic (4.4%), injuries and poisonings (4.4%), symptoms (2.9%), dermatologic (1.5%), and musculoskeletal (1.5%). These groupings include a significant enrichment of *delayed milestones (P =* 1.63 × 10^−10^; β, 1.22; SE, 0.19), *lack of normal physiological development (P =* 6.52 × 10^−7^; β, 0.59; SE, 0.12), and *disturbance of skin sensation (P =* x 1.39 × 10^−8^; β, 1.44; SE, 0.25) in TD patients. TD controls are significantly enriched for common childhood ailments, including: *acute upper respiratory infections of multiple or unspecified* sites *(P =* 4.91 × 10^−9^; β, -0.57; SE, 0.10), *fever of unknown origin (P =* 4.22 × 10^−8^; β, -0.72; SE, 0.13), *fracture in upper limb (P =* 9.36 × 10^−6^; β, -0.68; SE, 0.15), *fracture in hand or wrist* (*P* = 2.15 × 10^−5^; β, -1.47; SE, 0.35), and *cough (P =* 3.21 × 10^−5^; β, -0.59; SE, 0.14) (**Supplemental Table 2**), consistent with the fact that our TD control population were age-matched to TD cases, and consist of predominantly young individuals attending the health center for common childhood ailments (average age = 24.25 years, **Table 1**). There is a significant depletion of digestive and sense organ phenotypes in our TD findings *(P =* 1.8 × 10^−3^ and *P* = 2.0 × 10^−3^, respectively). Overall, our TD diagnosis PheWAS findings are congruent with established clinical features of TD and reveal the complex phenotypic architecture within individuals diagnosed with tic disorders.

### TD PheRS identifies individuals with shared phenotypical features of tic disorder patients

To consolidate the complex medical phenome of TD into a single, quantitative measure, we constructed a phenotype risk score (PheRS) using the 69 phenotypic features identified by the TD PheWAS (**Supplemental Table 2, Figure 4A**). We identified the PheRS features in the non-genotyped population of the Vanderbilt SD (discovery sample) by performing the PheWAS described above. The significantly associated phenotypes were then used to calculate the PheRS in the genotyped BioVU population (target sample) (**Supplemental Table 7, Supplemental Table 8**). As expected, most patients in our target population (59.83%) have a very low TD PheRS because they do not exhibit any phenotypic features of TD (**Figure 4B**).

**Figure 4:**
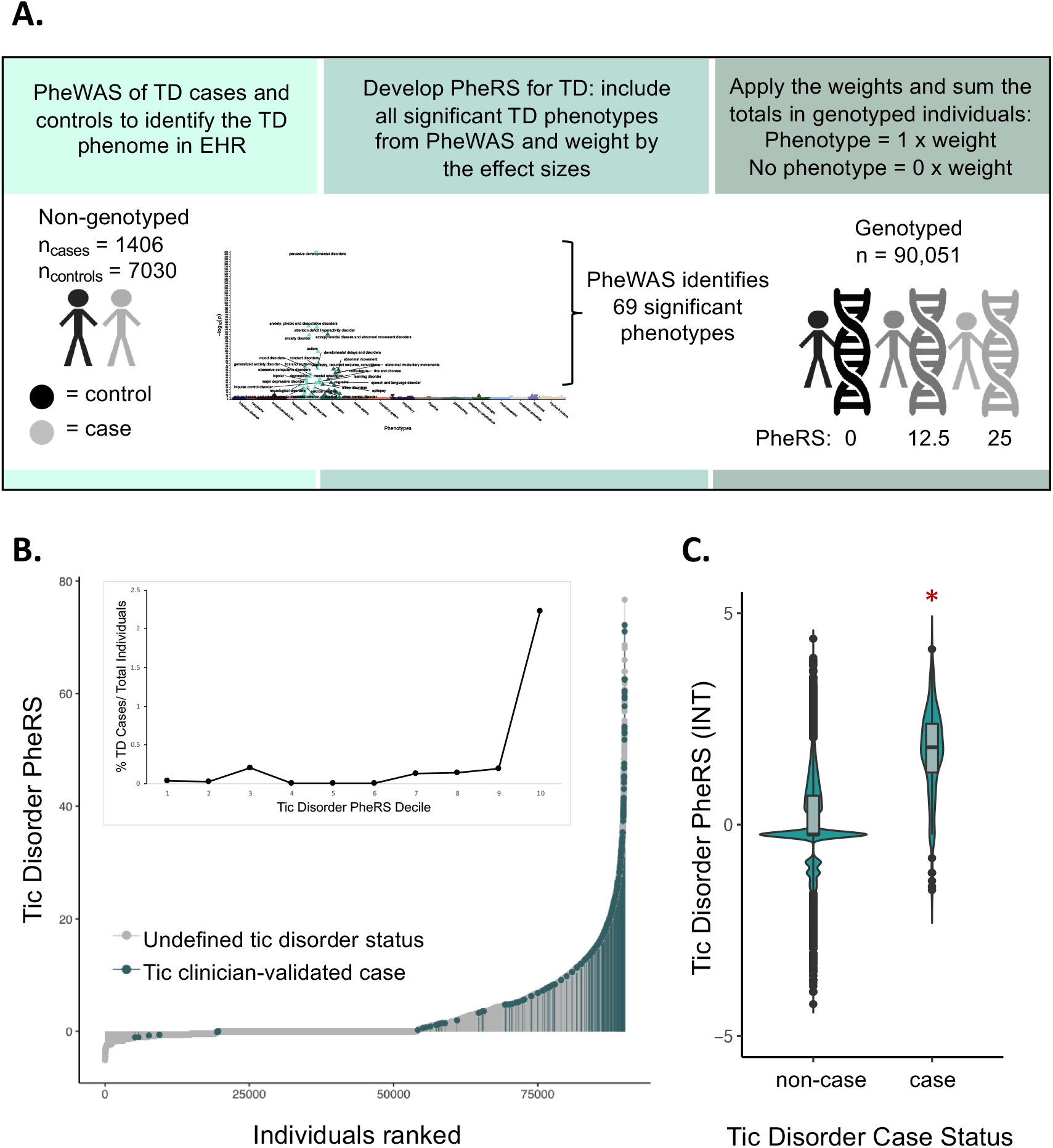
TD Phenotype Risk Score (PheRS) identifies individuals with shared features of tic disorders. A. The TD PheRS was calculated for 90,051 individuals in the Vanderbilt biobank using the 69 phenotypes identified in the TD PheWAS. Each individual is given a 0 or 1 for the absence or presence of each phenotype and this value is multiplied by the weight of each phenotype (weight = effect estimate from PheWAS analysis). B. The distribution of the TD PheRS across all genotyped individuals with clinician-validated cases highlighted in blue. All individuals not identified as cases by the TD algorithm and clinician review are gray. Inset shows the percentage of clinician-validated cases within each TD PheRS decile. C. Violin plots with boxplot insets of the TD PheRS in clinician-validated cases and non-cases shows that the TD PheRS in cases is significantly higher compared to controls (Wilcoxon rank sum test, *P*< 2.2 × 10^−16^, logistic regression analysis *P* = 4.787 × 10^−151^). The TD PheRS was inverse-normal transformed (INT) before plotting. In the logistic regression analysis, which was performed in the individuals of European ancestry, the covariates included PC1-10, age, median age of medical record, number of medical center visits, and sex.

Across the target population, we find that individuals of European ancestry (EA) have a significantly higher mean TD PheRS compared to individuals of African ancestry (AA); however, the EA sample size is much larger which may contribute to this finding *(P* < 2 × 10^−16^; β, 0.1; SE, 8.75 × 10^−3^; N_EA/AA_ = 70,439/15,174). We further find that the TD PheRS is significantly higher in females compared to males across the BioVU population *(P*=6.02 × 10^−16^, β, -0.05; SE, 6.6 × 10^−3^; N_female/male_ = 51,183/38,865). The TD PheRS is also inversely associated with current age and median age of medical record *(P<* 2 × 10^−16^; β, -4.0 × 10^−3^; SE, 1.0 × 10^−4^), suggesting that younger individuals are more likely to receive codes that contribute to the TD PheRS. Despite being younger, we find that individuals with a higher TD PheRS also have a greater number of visits to the medical center *(P<* 2.0 × 10^−16^; β, 5.38 × 10^−3^; SE, 3.98 × 10^−5^, **Supplemental Table 9**). This finding is expected, as the TD PheRS calculation requires accumulation of phecodes during visits to the medical center.

### TD PheRS is significantly higher in clinically-validated tic cases

Within the BioVU population, we identified by the presence of a tic disorder ICD9/10 diagnosis code or a mention of specific tic disorder keywords (**Supplemental Figure 1**). This approach identified 485 putative tic disorder cases, 316 of these were in the target sample. After clinician chart review of these 316 charts, 266 were confirmed as a gold-standard clinician reviewed tic disorder sample (84.2%).

The gold-standard set then served as a positive control for the TD PheRS (**Supplemental Tables 10 and 11**). We found that 75.6% (201/266) of the TD clinician-defined cases had PheRS scores within the tenth decile of the TD PheRS, and 91.4% (243/266) of the cases had PheRS scores falling in the seventh PheRS decile or higher (**Figure 4B**). Additionally, we find that clinically validated cases have a significantly higher TD PheRS compared to non-cases according to the Wilcoxon rank-sum test *(P<* 2.2 × 10^−16^) and logistic regression analysis accounting for covariates (*P* = 4.787 × 10^−151^; β, 1.68; SE, 0.06; **Figure 4C**).

A small proportion of the gold-standard set (23/266, 8.6%) have a very low TD PheRS, falling in the first three deciles (**Figure 4B**). Of the 23 confirmed TD cases with a PheRS equal to zero or less, 6 did not meet the medical home criteria despite having mentions of a TD diagnosis in their medical records, making these cases difficult to identify without the use of natural language processing algorithms. For the remaining 17 individuals, despite frequent visits to the medical center and documentation of TD in the clinical notes, the TD phenotype was never coded. These individuals have no ICD9/10 codes in common with tic disorder patients, but have mentions of Tourette’s, tics, chronic motor tic disorder, vocal and motor tics, or head tics in their medical records, indicating that these individuals likely visited the medical center for care independent of their tics, but included discussion of tics during their clinical encounter. Because the TD PheRS relies solely on the presence or absence of ICD9/10 billing codes, these individuals have a much lower PheRS despite having a TD diagnosis. Conversely, there are multiple BioVU patients with a high TD PheRS that were not identified by the TD algorithm and clinician validation (825 individuals within the top percentile, **Figure 4B**). These individuals share substantial overlapping phenome with TD patients, despite few of them having a TD phecode (15.3%, 126/825). Within the top percentile of the TD PheRS, over 50% of patients have a broad range of diagnoses including: malaise and fatigue, abdominal pain, anxiety disorder, convulsions, nausea and vomiting, depression, major depressive disorder, mood disorders, gastroesophageal reflux disease (GERD), and cough (**Supplemental Table 12**).

## Discussion

The availability of de-identified medical records for research purposes provides an expedited and relatively low-cost way to study phenotypically complex disorders over the lifespan in large patient populations. In this study, we leveraged the EHR database at Vanderbilt to build the PheRS, and the clinical biobank, BioVU, to evaluate its performance. We identified TD cases using ICD9/10 diagnosis codes, matched TD controls, and tested 676 phenotypes within the EHR for enrichment or depletion among TD cases. Consistent with prior studies, we found complex overlapping phenome between tic disorders and several neuropsychiatric phenotypes, with tic disorders most often being the first neuropsychiatric diagnosis in the patients’ medical records (∼64%), consistent with the high rate of community referrals to Vanderbilt Neurology. Additionally, we see that 51.9% of individuals diagnosed with tic disorders received at least two additional psychiatric diagnoses later in life, with 3.1 additional diagnoses on average. Even after conditioning on tic disorder medication use, we find a high prevalence of anxiety disorders (29.6%), attention-deficit hyperactivity disorder (19.1%), mood disorders (16.9%), depression (10.7%), autism (9.4%), and obsessive-compulsive disorder (8.7%) in individuals diagnosed with tic disorders, which may negatively impact these patients more severely than the tics themselves (**Supplemental Table 3, Supplemental Table 5**). These findings emphasize the complex phenotypic trajectories for tic patients and support the need for thorough observation and follow-up by clinicians and caregivers even if tic symptoms subside.

Phenotype risk scores can be used in research to quantify the phenomic patterns of comorbidity that characterize tic disorders. This single quantitative score can then be used to investigate broader phenotypic, and even genetic, liability to tic disorders. In this proof of principle study, we show that the TD PheRS is significantly higher in confirmed TD cases. Indeed, three quarters of the clinically validated TD cases had a PheRS in the highest decile. Additionally, there were several BioVU individuals in the top percentile of the TD PheRS lacking any TD phecode within their medical record (699/825, 84.7%). These may represent true TD patients missing a diagnosis code for TD or may represent individuals with several features of TD, such as the several neuropsychiatric comorbidities, without the presence of tics. These findings represent the strength of the PheRS approach at identifying individuals that would have otherwise been labeled as controls and may reduce power in a case-control study because they share many common features with TD patients. The strength of the PheRS lies in the ability to identify individuals with many features of a disease but without a formal diagnosis annotated in the EHR. Additional research is needed to determine how informative PheRS phenotypes will be for additional epidemiological and genetic applications.

Ultimately, we find that leveraging the dense phenotype information in a large, clinical biobank can replicate the phenotypic findings of prior studies of tic disorder patients, while adding longitudinal medical outcomes. In addition to the utility of the biobank for research purposes, these tools may also benefit clinicians and TD patients. Our PheWAS reveals multiple comorbid psychiatric phenotypes with tic disorders which could help clinicians more thoroughly evaluate TD patients and provide necessary interventions and treatments. Providing TD patients with a deeper understanding of their long-term risks of psychiatric disorders may allow the patients themselves to identify their symptoms and seek medical attention earlier. The PheRS serves as a quantitative measure encapsulating the broad medical phenome for a given disease. Because the PheRS is diagnosis agnostic and can be applied to target populations with any sample size, this tool could be advantageous for evaluating disease risk based purely on the presentation of phenotypes, and provides a phenome-wide perspective when evaluating complex disorders.

### Limitations

Despite the many conveniences of exploring phenotype information from the EHR, there are substantial limitations. The PheWAS is inherently biased by the number of medical center visits and number of ICD9/10 billing codes collected during these visits. A missing diagnosis code does not necessarily reflect an individual without the disease of interest and could be the result of a patient not receiving all of their care at a single hospital system, true cases visiting the hospital system for care unrelated to the disease of interest, or miscoding by the medical center. For these reasons, running case-control studies based on a single diagnosis code can be skewed by misclassification of case and control status. In our approach we have attempted to correct for some of these biases by limiting the controls of our TD PheWAS to the “medical home” population, requiring 5 diagnosis codes of any type on separate days over three consecutive years, to enrich for individuals that receive their primary care at Vanderbilt. Furthermore, the PheRS approach can alleviate some of the EHR coding biases because the score is calculated across several phenotypes to identify patients that share multiple, distinct phenotypic features with the designated case population. It is also likely that the individuals diagnosed with TD at Vanderbilt represent more severely affected cases due to ascertainment bias of a tertiary care center.

## Supporting information

Supplemental Methods and Results

## Data Availability

Because of patient confidentiality, individual-level data from BioVU and the Synthetic Derivative at Vanderbilt University Medical Center cannot be shared as it includes comprehensive electronic health record information. Data is available upon request and approval from Vanderbilt Institute for Clinical and Translational Research for researchers who meet the criteria for access to this confidential dataset. The phenotypes used to construct the tic disorder phenotype risk scores and the summary-level results are able to be shared and this information is included in the supplemental files of this manuscript.

## Conclusions

We demonstrate the complex clinical phenome that underlies TD diagnosis. Additionally, we validate a TD PheRS with the capacity to quantify tic risk based only on the phenotypic features captured in the EHR. With the increasing popularity of EHR usage in research, it may be possible to leverage the PheRS to increase the sample sizes and fidelity of TD cases and controls for future analyses.

## Acknowledgements

The authors acknowledge helpful discussions during the development of the tic disorder phenotype risk score and EHR algorithm, provided by the Psychiatric Genetic Consortium Tourette Syndrome Workgroup and the Tourette Syndrome Workgroup at Vanderbilt University Medical Center. The Synthetic Derivative and BioVU projects at VUMC are supported by numerous sources: including the NIH funded Shared Instrumentation Grant S10OD017985 and S10RR025141; CTSA grants UL1TR002243, UL1TR000445, and UL1RR024975 from the National Center for Advancing Translational Sciences. Its contents are solely the responsibility of the authors and do not necessarily represent official views of the National Center for Advancing Translational Sciences or the National Institutes of Health. Genomic data are also supported by investigator-led projects that include U01HG004798, R01NS032830, RC2GM092618, P50GM115305, U01HG006378, U19HL065962, R01HD074711; and additional funding sources listed at https://victr.vumc.org/biovu-funding/.

