## Supplemental Methods and Results for "Developing a Phenotype Risk Score for Tic Disorders in a Large, Clinical Biobank"

**The Synthetic Derivative (SD) is a database of de-identified electronic health records (EHRs)**

The SD houses clinical information and documentation for over 3.6 million individuals who receive clinical care at Vanderbilt University Medical Center (VUMC) dating back to 1994. This information includes insurance billing codes (International Classification of Diseases, 9$^{th}$ and 10$^{th}$ editions/ICD-9 and ICD-10 codes, respectively), clinical procedure codes (Current Procedural Terminology/CPT codes), clinician notes, family histories, lab values, and prescribed medications[1].

**BioVU is a biorepository of genotype data linked to medical records**

The Vanderbilt Institute for Clinical and Translational Research at VUMC curates BioVU, a clinical biorepository linked to the de-identified EHR information within the SD[2]. Patients seen at a Vanderbilt clinic are given the option to participate in the BioVU research program, which collects the leftover blood samples from routine clinical testing for genotyping and research. Sample collection for BioVU began in 2007 and is ongoing at Vanderbilt clinics across middle Tennessee. Currently, the BioVU biobank houses DNA samples linked to de-identified EHRs for 298,378 individuals.

**Identification of TD cases and controls in the EHR**

We identified TD cases and controls within the EHR using the following criteria. TD cases were required to have at least two separate, temporally distinct instances of case inclusion TD phenotypes, defined by ICD-9 and ICD-10 (International Classification of diseases, Ninth/Tenth Revision) billing codes (**Supplemental Table 1**). To prevent individuals with confounding phenotypes from being included in the case group, potential TD cases were excluded from the analysis if any instance of TD exclusion codes (such as non-tic movement disorders) were present in the medical record (**Supplemental Table 1**). As the genotyped individuals served as the validation sample, TD cases were restricted to the non-genotyped population within the synthetic derivative (SD) to yield 1,406 individuals. Controls were selected from the medical home population within the SD, individuals that had visited a Vanderbilt clinic at least five times within a consecutive three-year period. TD controls were restricted to the non-genotyped population within the synthetic derivative and were age and sex matched to the TD cases (5:1) using the MatchIt package in R[3]. TD controls were excluded if any instances of either the TD inclusion or exclusion codes were present in the medical record, resulting in 7,030 TD controls. To test for significant differences in demographic values between TD cases and controls, an independent-samples t-test was performed for each variable between cases and controls. Chi-squared tests were performed to evaluate differences in proportions of ethnicity and EHR-reported race between cases and controls (**Table 1**).

**PheWAS Sensitivity Analysis**

In a sensitivity analysis, we conditioned the TD PheWAS on the presence or absence of commonly-prescribed TD medications from the medical records (i.e. risperidone, aripiprazole, fluphenazine, haloperidol, guanfacine, and clonidine).

**Algorithm and clinician validation of TD cases in BioVU**

For prior genetic analyses, we sought to identify BioVU individuals with a clear clinical diagnosis of TD. A combination of tic disorder diagnosis codes and keywords were used to define a TD algorithm. Inclusion criteria required at least two instances of tic disorder ICD-9 billing codes within the EHR (**Supplemental Figure 1**) or the single presence of the keywords *motor tic, vocal tic, Tourette,* or *tic disorder* in the clinical notes. Exclusion criteria included ICD-9 billing codes for muscular diseases (**Supplemental Figure 1**). The tic disorder algorithm identified 485 cases, 408 of which subsequently underwent clinician chart review. Of the 408 algorithm cases, 316 were within the BioVU dataset and 266 BioVU individuals were clinically-validated as true tic disorder cases (84.2%). The patients identified by the TD algorithm that were not confirmed by clinician chart review were excluded based on multiple criteria (i.e. having a broad mention of tics in the medical history without additional information, tics that appeared following medication use, and mentions of tic misdiagnosis after appearance of abnormal movements or seizures). Of the 266 clinically-validated TD individuals, 95 met the ICD9/10 code criteria, 40 met the keyword criteria, while 131 met both.

### Supplemental Results

#### Tic disorders and abnormal movement phenotypes

The neurological phenotypes enriched in TD cases (*extrapyramidal disease and abnormal movement disorders* ($P$= 1.11 x $10^{-135}$; β, 5.10; SE, 0.21), *abnormal movement* ($P$= 4.58 x $10^{-62}$; β, 2.00; SE, 0.12), *torsion dystonia* ($P$= 6.82 x $10^{-20}$; β, 3.43; SE, 0.38)) may represent misdiagnoses or the accumulation of diagnoses during a diagnostic odyssey. Abnormal movements can also occur in tandem with TD, as a secondary response to TD medication, or independently of TD[4]. To further investigate this, we performed a sensitivity analysis, conditioning the TD PheWAS on the presence or absence of commonly-prescribed medications for tics. Associations between hyperkinetic movement disorders and tic disorder diagnosis status remain after correcting for medications, suggesting that these movement phenotype associations are not solely side effects of TD medication usage (**Supplemental Table 5),** consistent with prior studies finding that nearly 10% of Tourette syndrome patients have co-occurring dystonia.[5,6] We find that within the EHR-derived TD cases, 54% of individuals with a hyperkinetic movement diagnosis received a tic disorder diagnosis first, while the remaining 46% received a movement diagnosis first; however, it is difficult to disentangle these findings, which cannot rule out multiple possibilities for the co-occurring codes, including clinical misdiagnoses (**Supplemental Table 6**)[4].

**Supplemental Figures:**

A.

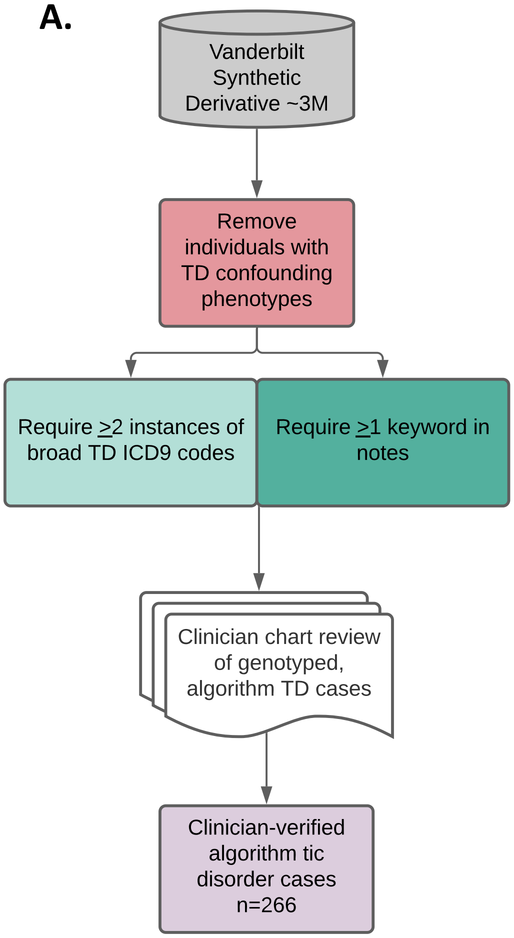


B.

| Tic Disorder (TD) Confounding ICD-9 Codes | |
|---|---|
| ICD-9 Code | Phenotype |
| 333.6 | Genetic torsion dystonia |
| 333.7 | Acquired torsion dystonia |
| 333.72 | Acute dystonia due to drugs |
| 333.79 | Other acquired torsion dystonia |
| 333.8 | Fragments of torsion dystonia |
| 333.82 | Orafacial dyskinesia |
| 333.85 | Subacute dyskinesia due to drugs |
| 333.89 | Other fragments of torsion dystonia |

C.

| Broad Tic Disorder (TD) ICD-9 Codes | |
|---|---|
| ICD-9 Code | Phenotype |
| 307.2 | Tics |
| 307.20 | Tic disorder NOS |
| 307.21 | Transient tic disorder |
| 307.22 | Chronic motor or vocal tic disorder |
| 307.23 | Tourette's disorder |
| 333.3 | Tics of organic origin |

D.

| Tic Disorder (TD) keywords |
|---|
| motor tic |
| vocal tic |
| Tourette |
| tic disorder |

**Supplemental Figure 1: TD algorithm and clinician chart review identify tic disorder cases in the EHR.** A. The Synthetic derivative (SD) at Vanderbilt consists of de-identified electronic health records for ~3 million individuals. To identify individuals with tic disorders and exclude individuals with confounding phenotypes, patients with torsion dystonia and dyskinesia diagnoses were excluded. The TD algorithm required at least 2 instances of tic disorder ICD-9 codes and 1 tic disorder keyword in the records for each individual. When restricted to genotyped individuals, this algorithm identified 408 TD cases. Clinician chart review was performed on the 408 individuals, which resulted in 266 clinician-validated TD cases.  B. Table of TD-confounding phenotypes which were used as exclusion criteria in the algorithm. C. Table of TD ICD-9 codes which were used to identify TD cases. D. Table of TD keywords which were used to identify TD cases.

**Supplemental Table Legends:**

**Supplemental Table 1: Tic Disorder (TD) case and control inclusion and exclusion criteria.** To define the TD cases for PheWAS, at least two instances of ICD9/10 billing codes for broad tic disorders were required. Cases were excluded if they had any mention of the ICD9/10 exclusion codes and cases were filtered for non-genotyped individuals. Requiring 2 instances of a code removed 2,454 individuals, requiring no instances of the exclusion criteria removed 112 individuals, and subsetting to the non-genotyped population removed 134 individuals. Controls were excluded if they have any mention of the ICD9/10 inclusion or exclusion codes.

**Supplemental Table 2: Tic Disorder PheWAS results.** A phenome-wide association study was performed across TD cases (N=1,406) and matched controls (N=7,030). 69 of the 676 phenotypes tested were significantly associated with TD case status after Bonferroni correction ($P = 7.396 \times 10^{-5}$). Covariates included: current age, sex, self-reported race, ethnicity, median age within medical record, and number of visits to medical center. Abbreviations: SE (standard error), OR (odds ratio).

**Supplemental Table 3: Tic Disorder Psychiatric Comorbidities.** The TD PheWAS results included significant associations between several psychiatric phenotypes with TD case status.

This table includes a breakdown of each psychiatric phecode with the number and percent of TD cases (N=1,406) and TD controls (7,030) that also have each psychiatric diagnosis.

**Supplemental Table 4: First Neuropsychiatric Diagnosis for TD Cases.** For the 1,406 TD cases, we extracted the first psychiatric diagnosis from each TD individual's medical record to examine whether TD cases are typically diagnosed with tic disorders before receiving diagnoses for other psychiatric phenotypes. This table lists the counts and percent of individuals that have a first diagnosis of each listed psychiatric phenotype.

**Supplemental Table 5: TD PheWAS Results after Conditioning on TD Medications.** The TD PheWAS was conditioned by the absence or presence of commonly prescribed TD medications (see Methods) extracted from the medical records to determine whether the movement disorder phenotype associations were driven by TD medication use. The PheWAS was performed as before, with TD cases (N=1,406) and matched controls (N=7,030). 49 of the 676 phenotypes tested were significantly associated with TD case status after Bonferroni correction ($P$= 7.396 x $10^{-5}$). Covariates included: current age, sex, EHR-reported race, EHR-reported ethnicity, median age within medical record, number of visits to medical center, and presence/absence of TD medication. Abbreviations: SD (standard deviation), OR (odds ratio).

**Supplemental Table 6: First Diagnosis for TD Cases with Hyperkinetic Movement Diagnosis.** For the 185 TD cases with a hyperkinetic movement disorder diagnosis, we extracted the first diagnosis of psychiatric or movement disorders from each individual's medical record.

This table lists the counts and percent of individuals that have a first diagnosis of each listed phenotype.

**Supplemental Table 7: BioVU demographics.** The BioVU MEGA cohort consists of 90,051 individuals with genetic data linked to de-identified electronic health records. The phenotype risk score for tic disorders was generated in the non-genotyped individuals and applied to this independent population by utilizing the available phenotype data.

**Supplemental Table 8: TD PheRS Phenotypes and Weights.** The 69 phenotypes significantly associated with TD case status were used to generate the TD PheRS. Each phenotype is weighted by the effect size estimate (β) calculated from the phenome-wide association study.

**Supplemental Table 9: TD PheRS Statistical Tests in BioVU Participants.** Linear and logistic regression models were used to calculate the relationships between the TD PheRS within individuals of European or African ancestry, across sex, current age, median age of medical record, number of medical center visits, and presence or absence of tic disorder phecode. A higher TD PheRS is associated with European ancestry, female sex, younger current age, younger median age of medical record, higher numbers of medical center visits, and the presence of a tic disorder phecode.

**Supplemental Table 10: Clinician-validated TD cohort demographics.** A combination of ICD-9 billing codes and keywords within the medical records were utilized to identify patients diagnosed with TD. Of the 408 individuals identified by the algorithm, 266 were confirmed TD cases upon clinician chart review.

**Supplemental Table 11: TD PheRS phenome in the clinician-validated TD Individuals.** 69 phenotypes were used to calculate the TD PheRS. These phenotypes were represented in the individuals identified as TD cases by the TD algorithm and clinician review. The tics and stuttering phenotype was the most commonly observed phenotype, found in over 50% of the TD clinician-validated cases.

**Supplemental Table 12: All clinical phenome represented in the TD PheRS top percentile.** All of the phenotypes represented within the BioVU participants in the top percentile of the TD PheRS (total N = 901).
